# Computational analysis of quantitative echocardiographic assessments of functional mitral regurgitation: Proximal Isovelocity Surface Area (PISA) methods

**DOI:** 10.1101/2021.09.28.21264279

**Authors:** Tongran Qin, Andrés Caballero, Rebecca T. Hahn, Raymond McKay, Wei Sun

**Affiliations:** Tissue Mechanics Laboratory, The Wallace H. Coulter Department of Biomedical Engineering, Georgia Institute of Technology and Emory University, Atlanta, GA, USA; Division of Cardiology, Columbia University Medical Center, New York, NY, USA; Cardiology Department, The Hartford Hospital, Hartford, CT, USA

**Keywords:** proximal isovelocity surface area (PISA), integrated PISA, echocardiography, EROA, regurgitant volume, mitral regurgitation

## Abstract

While proximal isovelocity surface area (PISA) method is one of the most common echocardiographic methods for quantitative mitral regurgitation (MR) assessment, accurate MR quantification remains challenging. This study examined the theoretical background of PISA, performed virtual echocardiography on computer models of functional MR, and quantified different sources of errors in PISA. For regurgitant flow rate measurement, the conventional 2D hemispherical PISA caused significant underestimation due to underestimation of PISA area, the multiplane 2D hemiellipsoidal and hemicylindrical PISA provided improved accuracy with better assumptions on PISA contour shape. With the direct capture of PISA area, the 3D-PISA was found to be the most accurate. However, it should be noted that PISA method is subject to systematic underestimation due to the Doppler angle effect, and systematic overestimation due to the “flow direction angle” between the regurgitant flow direction and the PISA contour normal direction. For regurgitant volume quantification, integrated PISA, when performed properly, was able to capture the dynamic MR and therefore was more accurate than peak PISA. In specific, integrated PISA using the sum of regurgitant flow rates is recommended.

**Objectives:** The aim of this study was to evaluate the accuracy of different proximal isovelocity surface area (PISA) methods, examine their theoretical background, and quantify multiple sources of error in functional mitral regurgitation (MR) assessment.

**Background:** While PISA method is one of the most common echocardiographic methods for MR severity assessment, it is associated with multiple sources of errors, and accurate MR quantification remains challenging.

**Methods:** Five functional MR (FMR) computer models were created, validated and treated as phantom models. The phantom models have fully resolved and detailed flow fields in the left atrium (LA), left ventricle (LV) and cross the mitral valve, from which the reference values of mitral regurgitant flow rate and regurgitant volume can be obtained. The virtual PISA measurements (i.e., 3D and 2D PISA) were performed on the phantom models assuming optimal echo probe angulation and positioning. The results of different PISA methods were compared with the reference values.

**Results:** For regurgitant flow rate measurements, compared to the reference values, excellent correlations were observed for 3D-PISA (R = 0.97, bias -24.4 ± 55.5 ml/s), followed by multiplane 2D hemicylindrical (HC)-PISA (R = 0.88, bias -24.1 ± 85.4 ml/s) and hemiellipsoidal (HE)-PISA (R = 0.91, bias -55.7 ± 96.6 ml/s), while weaker correlations were observed for single plane 2D hemispherical (HS)-PISA with large underestimation (PLAX view: R = 0.71, bias -77.6 ± 124.5 ml/s; A2Ch view: R = 0.69, bias -52.0 ± 122.0 ml/s; A4Ch view: R = 0.82, bias -65.5 ± 107.3 ml/s). For regurgitant volume (RV) quantification, integrated PISA presented improved accuracy over peak PISA for all PISA methods. For 3D-PISA, the bias in RV improved from -12.7 ± 7.8 ml (peak PISA) to -2.1 ± 5.3 ml (integrated PISA).

**Conclusions:** In FMR, conventional single plane 2D HS-PISA significantly underestimated MR, multiplane 2D PISA (HE-PISA and HC-PISA) improved the accuracy, and 3D-PISA is the most accurate. To better capture the dynamic feature of MR, integrated PISA using the sum of regurgitant flow rates is recommended.

## INTRODUCTION

Mitral regurgitation (MR) is the most common valvular heart disease, with a prevalence of 9.3% in the US population aged 75 and above [1]. Although Doppler echocardiography (echo) is the primary clinical tool to evaluate MR severity, MR quantification remains challenging, and a true gold standard technique is still lacking [2, 3]. The proximal isovelocity surface area (PISA) method, as the most commonly used method for MR quantification, is derived from an ideal condition of the flow through an infinitesimal orifice on a flat plane [4], where the velocity is always perpendicular to well-defined hemispherical isovelocity shells. However, there are multiple sources of errors in practice when the ideal condition is not met, especially in functional MR (FMR). For the quantification of regurgitant flow rate, MR orifice in FMR is quite diverse [5], and is often elongated elliptical or crescent along the leaflet coaptation line [6], therefore, the PISA contour often deviates from hemisphere, and the flow velocity is not guaranteed to be perpendicular to the PISA contour. For the quantification of regurgitant volume (RV), the effective regurgitant orifice area (EROA) obtained at a single time point ignores the dynamic variation of regurgitant orifice [7-9]. All these factors could cause significant errors in the assessment of FMR.

Although many variations of PISA method have been proposed to account for the non-circular orifice (multiplane 2D-PISA [10-13], 3D-PISA [14-17]) and/or the dynamic variation (serial PISA [7], integrated PISA [18], mean EROA [19]) in FMR, there still lacks an in-depth analysis based on the fundamental theoretical background of PISA, which could not only identify but also quantify different sources of errors in PISA. The clinical studies on PISA are often limited by the lack of a robust reference value (gold standard) for MR quantification, where various reference values (quantitative volumetric method [20], cardiac magnetic resonance imaging [17, 18], vena contracta area [15, 16]) have been used. *Ex vivo* bench experimental studies are also challenging due to the complexity of experimental setups (e.g., beating heart conditions) and the difficulty to obtain the detailed interrogation of the MR flow field crossing the mitral valve (MV). Recent advances in patient image-based computational modeling, on the other hand, have substantially enhanced the investigation of patient-specific cardiac dynamics. In particular, patient specific finite element (FE) and fluid-structure interaction (FSI) computer models has been applied to study valvular functions under a variety of physiologic, pathologic and post-procedure states [21-27], including the echo measurement for MR [24], where the computer models were able to replicate the patient-specific flow field inside the left heart. Although the flow field obtained from computer models might not exactly replicate the *in vivo* data due to simplifications and assumptions, the computer model itself can be used as a phantom model (similar to a bench-top experimental model) for systematical quantification of different sources of errors in echo measurements [24]. Therefore, in this study we will investigate the accuracy and reliability of PISA in FMR using computer models. From the computer models, fully resolved and detailed flow fields in the left atrium (LA), left ventricle (LV) and cross the MV will be extracted, from which the reference values of mitral regurgitation flow rate and regurgitant volume can be obtained. The virtual PISA measurements (i.e., 3D and 2D PISA) will be performed on the phantom models assuming optimal echo probe angulation and positioning. The results of different PISA methods will be compared with the reference values. The accuracy of different PISA methods will be evaluated, and the multiple sources of errors in PISA method will be analyzed and quantified.

## METHODS

### Computer models

In this study, five FMR computer models were created based on the FSI modeling framework developed by our group [21-25]. The first model was a previously validated patient-specific left heart FMR computer model based on a 71-year-old male patient [25] (data collected under the Institutional Review Broad approvals). The other four FMR models were computationally created from the first model by displacing the papillary muscles (PM) apically and laterally, replicating a type IIIb lesion with restricted leaflet motion.

The 3D flow field (Figure 1) and the flow rate across the aortic value (AV) and MV can be directly extracted from the computer models over the entire cardiac cycle, and treated as the reference values. Specifically, the reference MV regurgitant volume *RV*_*MV*_ was obtained by integrating the negative MV flow rate over systole. The regurgitant fraction *RF*_*MV*_ = *RV*_*MV*_/*Sv*_*LV*_ was used to grade MR severity, where *Sv*_*LV*_ = *Sv*_*AV*_ + *RV*_*MV*_ is the total stroke volume (SV) of the left ventricle (LV). Based on the *RF*_*MV*_ [3], Among the five FMR models, Patient 1 (P1) and P2 have mild-moderate MR, P3 has moderate-severe MR, while P4 and P5 have severe MR (Table 1).

**Figure 1.**
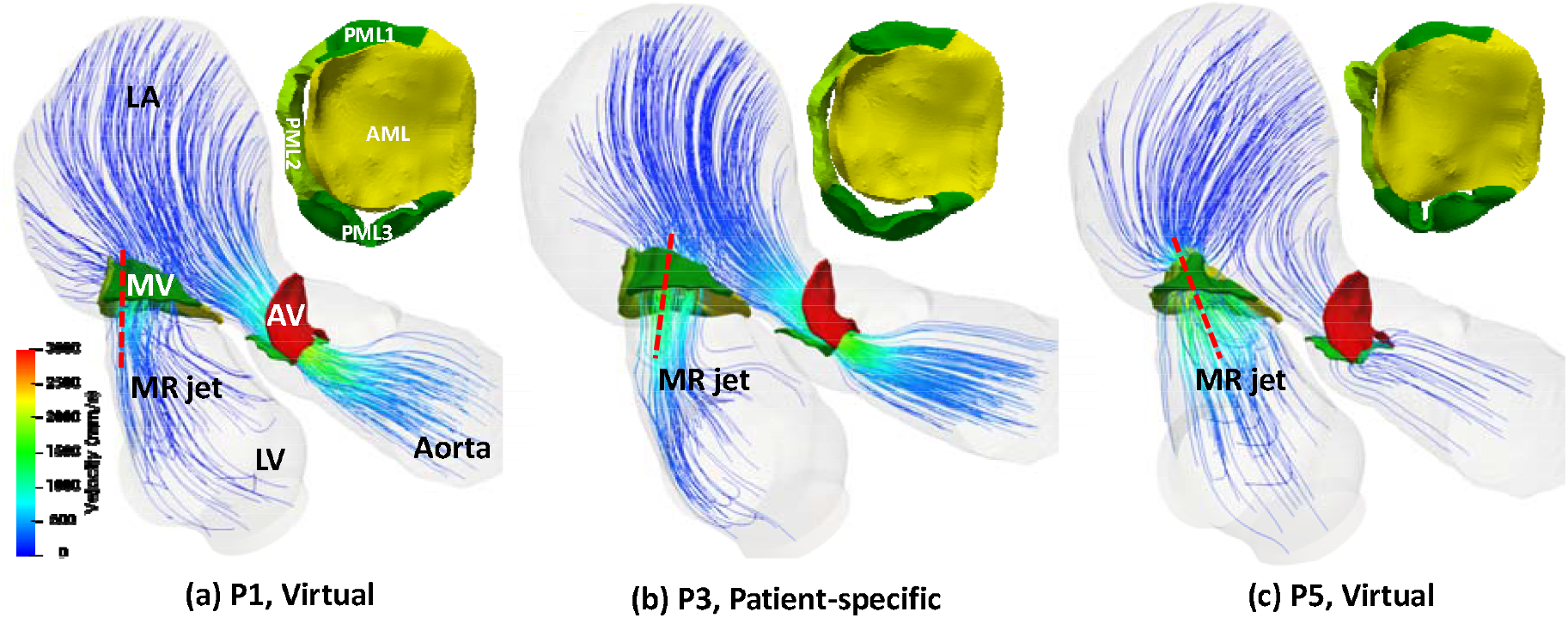
The velocity field of three representative FMR computer models (P1, P3, and P5) ranging from mild to severe MR, and the corresponding top (ventricular) view of MV at peak MR. The red dashed lines denote the direction of the MR jet.

**Table 1.** Patient (FMR computer model) information

| Patient (computer FMR model) | P1<br>Virtual | P2<br>Virtual | P3<br>Patient-specific | P4<br>Virtual | P5<br>Virtual |
| --- | --- | --- | --- | --- | --- |
| $SV_{AV}$ (ml) | 55.65 | 51.02 | 46.28 | 33.71 | 21.52 |
| $SV_{MV}$ (ml) | 74.59 | 75.58 | 74.64 | 77.26 | 75.61 |
| $RV_{MV}$ (ml) | 27.68 | 32.61 | 37.59 | 51.62 | 62.69 |
| $RF_{MV}$ (%) | 33.22 | 38.99 | 44.82 | 60.50 | 74.45 |
| MR Severity (Based on $RF_{MV}$) | MILD-MODERATE | MILD-MODERATE | MODRATE-SEVERE | SEVERE | SEVERE |

### Virtual Echocardiographic data acquisitions

In this study, the velocity field obtained directly from computer models (Figure 1) is referred to as the “true velocity”, while the velocity field used for virtual echocardiographic data acquisition is referred to as the “Doppler velocity”. Due to the Doppler angle effect [2, 28-30], the virtual echo only measures the velocity component aligned with the ultrasound beam. Therefore, the Doppler velocity field was obtained by projecting the true velocity field along the direction of the ultrasound beam, which is assumed to be in its optimal positioning and angulation, i.e., along the direction of MR jet (Figure 1). Despite a well-known limitation of Doppler echo imaging, the error due to the Doppler effect is very challenging to quantify using *ex vivo* models and nearly impossible using *in vivo* models. However, it can be easily quantified using our computer models by comparing the results obtained from the true velocity and the Doppler velocity.

### True-PISA and 3D-PISA

Without any assumptions of PISA shape (i.e., spherical or elliptical), the PISA contour of an arbitrary shape can be obtained directly from the velocity field using the aliasing velocity *V*_*a*_ ranging from 30 - 50 cm/s [3, 31]. The PISA obtained using the true velocity field is referred to as true-PISA, while the PISA obtained using the Doppler velocity field is referred to as 3D-PISA (Figure 2d), which is the one that clinical 3D echo will obtain. It should be noted that the isovelocity shells for both true-PISA and 3D-PISA were identified using the velocity magnitude, not the normal velocity component across the PISA. For true-PISA and 3D-PISA quantification, the PISA surface area *S*_*PISA*_ was quantified by triangulation of their individual isovelocity shell.

**Figure 2.**
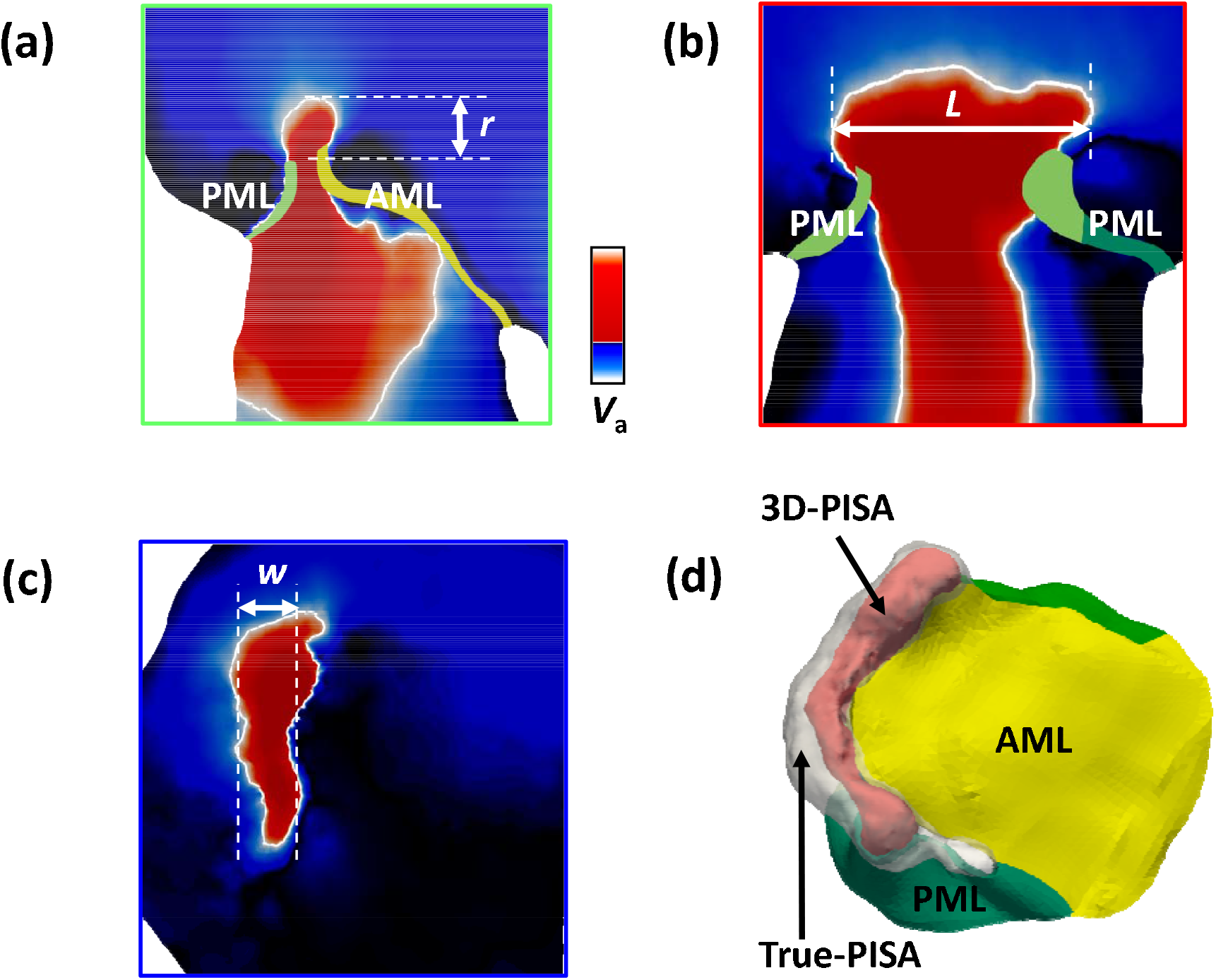
(a)-(c) The measurement of PISA radius *r*, length *L*, and width *w* in three orthogonal planes in HE-PISA and HC-PISA. White solid lines denote PISA contour. (d) The true-PISA contour (translucent white shell) based on the true velocity field, and the 3D-PISA contour (interior red shell) based on the Doppler velocity field.

### Single plane and multiplane 2D-PISA

The 2D-PISA contour area was calculated based on certain assumptions of PISA shape. In single plane 2D-PISA, the PISA contour was assumed *hemi-spherical (HS)*, and only one measurement of PISA radius *r* from a single plane is required, yielding the PISA area *S*_*PISA*_ = 2*πr*^2^. In this study, three different HS-PISA measurements were performed within different acoustic windows: the parasternal long-axis (PLAX) view, the apical 2-chamber (A2ch) view, and the apical 4-chamber (A4ch) view. In multiplane 2D-PISA, multiple dimensions of PISA contour were measured in orthogonal planes. Assuming a *hemiellipsoidal (HE)* shape, HE-PISA [10, 11] required measurements of the length *L*, the width *w*, and the radius *r* along the base of PISA (Figure 2a-c), yielding *S*_*PISA*_ = 2*π*[(*L*^*p*^*w*^*p*^ + *L*^*p*^*r*^*p*^ + *w*^*p*^*r*^*P*^)/3]^1/*p*^ [32], where *p* ≈ 1.6075. Assuming a *hemicylindrical (HC)* shape, HC-PISA [12, 13] required measurements of the length *L* and the radius *r* of PISA, yielding *S*_*PISA*_ = *πrL* + *πr*^2^.

### Quantification of MR

The regurgitant flow rate *Q*_*MR*_ = *V*_*a*_ × *S*_*PISA*_ is a product of the aliasing velocity *V*_*a*_ and the corresponding PISA contour area *S*_*PISA*_. The EROA and regurgitant volume *RV*_*MV*_ can then be calculated as *EROA* = *Q*_*MR*_ /*V*_*MR*_ and *RV*_*MV*_ = *EROA* × *VTI*_*MR*_, respectively. The regurgitant velocity *V*_*MR*_ and the velocity time integral *VTI*_*MR*_ of the regurgitant jet were obtained by emulating continuous wave Doppler (CWD) measurements with the virtual ultrasonic probe parallel to the jet in an optimal position.

Both peak PISA and integrated PISA measurements were performed. The peak PISA was obtained using *Q*_*MR*_ at one instance of peak MR, while the integrated PISA required measurements at multiple time frames over systole [33], with 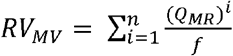, where 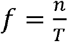 is the frame rate for PISA measurement, i.e., *n* is the total number of measured frames over systole (n=4 in this study), and T is the time interval of systole. The “integrated PISA” defined in this study is consistent to “serial PISA” in Ref.[7], and “integrated PISA” in Ref.[18]. For patients with multiple regurgitant jets, *RV*_*MV*_ was calculated separately for each jet, and summed over all jets.

### Statistical analysis

Continuous data are expressed as mean ± SD. Linear regression analysis was performed to evaluate the correlations between the values from different PISA methods and the reference MR values. Bland– Altman plots with limits of agreement were used to estimate the precision of different PISA methods, which showed the mean measurement bias ± 1.96 SDs. Interobserver and intraobserver reproducibility were evaluated using intraclass correlation coefficients. Differences were considered significant when P < .05 (two sided).

## RESULTS

### Measurement of the instantaneous regurgitant flow rate Q_MR_

Excellent correlations were observed in the *Q*_*MR*_ quantification between the true-PISA and reference value (R = 0.99, P <.001; Figure 3A), and between the 3D-PISA and reference value (R = 0.97, P <.001; Figure 3B). However, the regression equations and the Bland-Altman analysis indicated a consistent overestimation by true-PISA (bias 32.3 ± 35.2 ml/s, P <.001; Figure 3C), and a consistent underestimation by 3D-PISA (bias -24.4 ± 55.5 ml/s, P <.001; Figure 3D).

**Figure 3.**
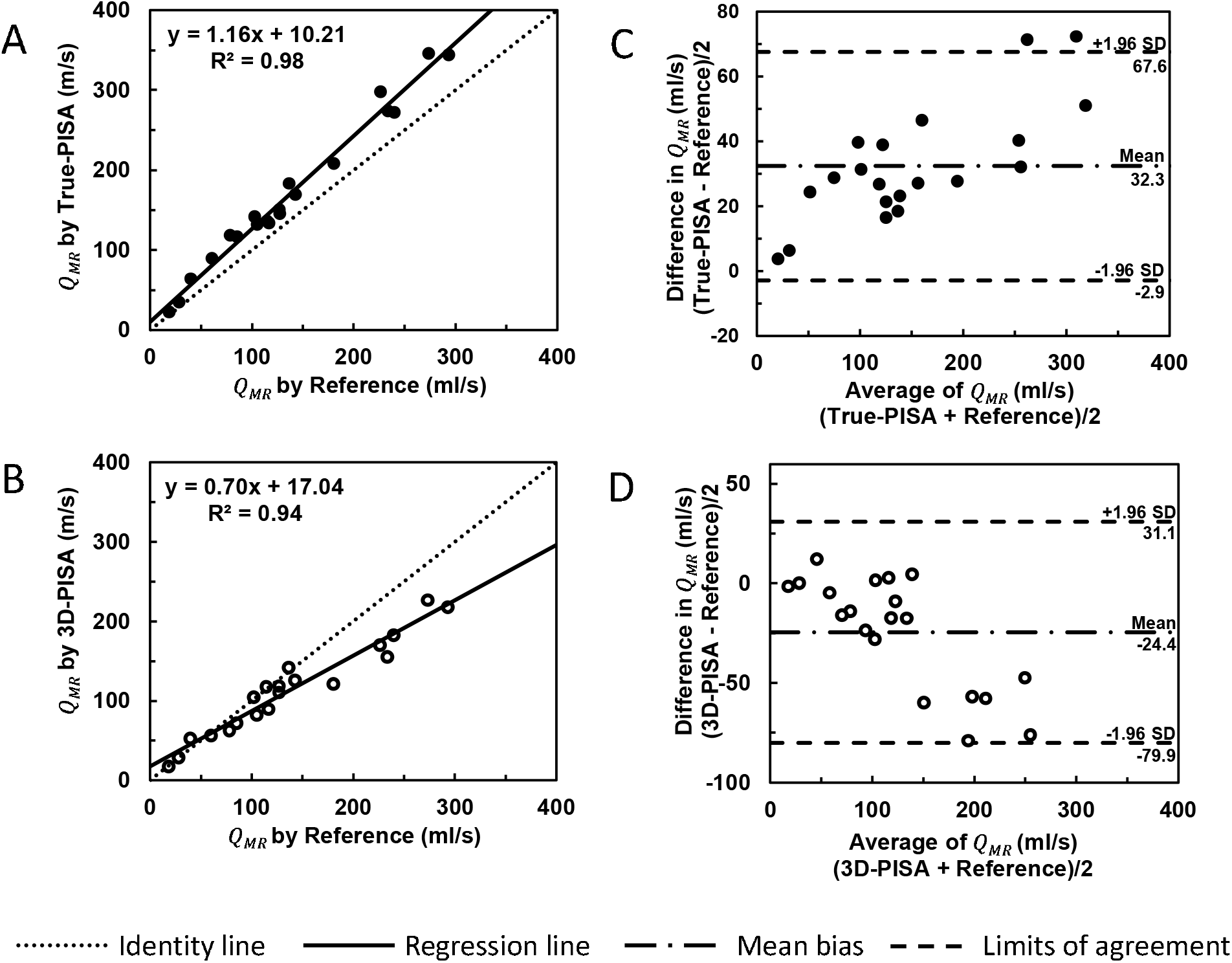
Linear regression plots showing correlations between *Q*_*MR*_ by reference, and *Q*_*MR*_ by true-PISA (A) and 3D-PISA (B). Bland-Altman plots showing the *Q*_*MR*_ measurements bias by true-PISA (C) and 3D-PISA (D). In Figures 3-5, different symbols represent the results obtained using different PISA methods. For each FMR model, four measurements were obtained over systole.

As seen in Figure 4, for both multiplane 2D PISA methods, good correlations were observed in the *Q*_*MR*_ quantification between the PISA method and reference value (For HC-PISA, R = 0.88, P <.001; Figure 4A; For HE-PISA, R = 0.91, P <.001; Figure 4B). The regression equations and the Bland-Altman analysis revealed a better *Q*_*MR*_ quantification by HC-PISA (bias -24.1 ± 85.4 ml/s, P <.001; Figure 4C) than by HE-PISA (bias -55.7 ± 96.6 ml/s, P <.001; Figure 4D).

**Figure 4.**
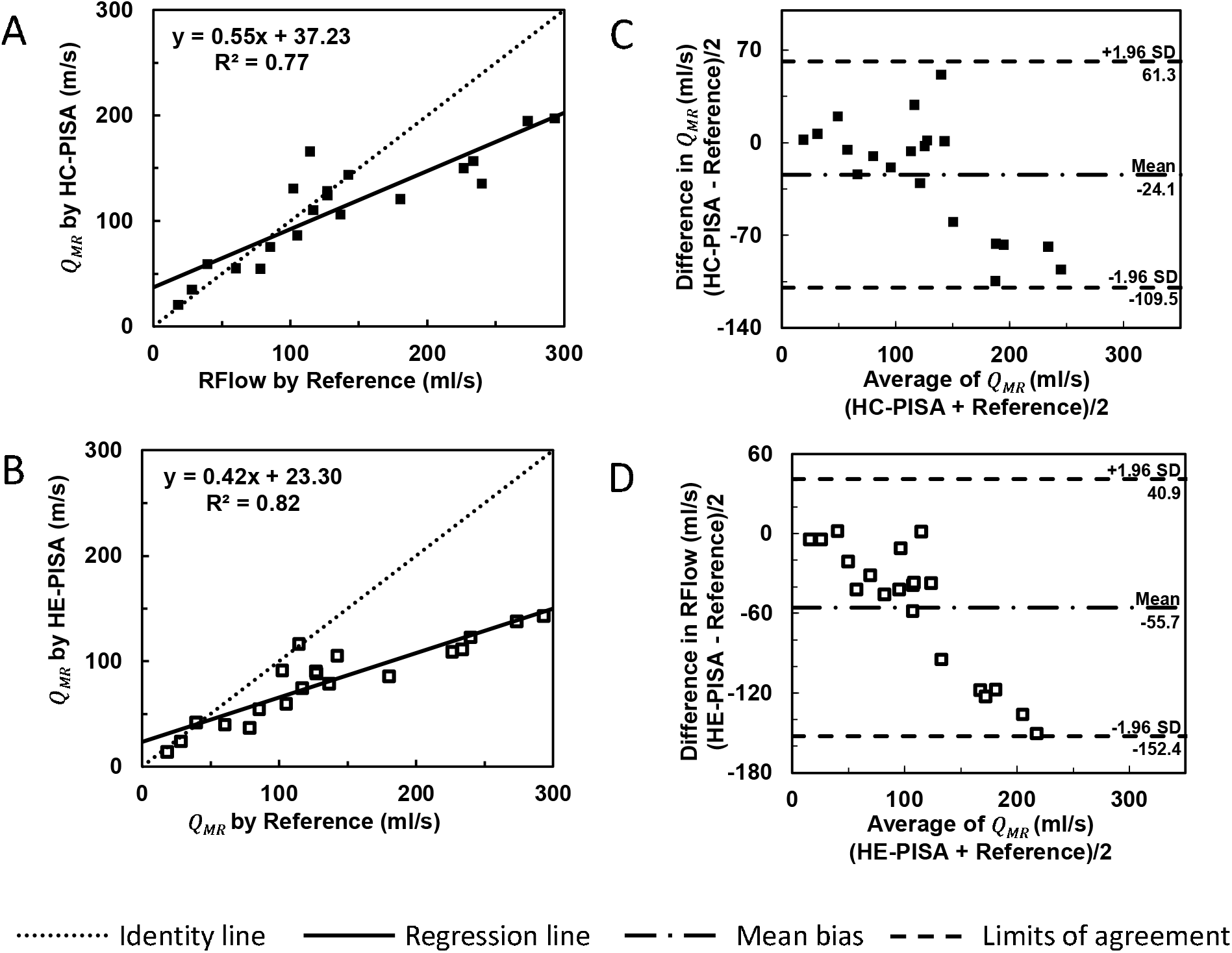
Linear regression plots showing correlations between *Q*_*MR*_ by reference, and *Q*_*MR*_ by HC-PISA (A) and HE-PISA (B). Bland-Altman plots showing the *Q*_*MR*_ measurements bias by HC-PISA (C) and HE-PISA (D).

For all single plane 2D HS-PISA, weaker correlations were observed in the *Q*_*MR*_ quantification between the PISA method and reference value (For PLAX HS-PISA, R = 0.71, P <.001; Figure 5A; For A2Ch HS-PISA, R = 0.69, P <.001; Figure 5B; For A4Ch HS-PISA, R = 0.82, P <.001; Figure 5C). The regression equations and the Bland-Altman analysis indicated large underestimations by all three HS-PISA methods, and the magnitude of *Q*_*MR*_ underestimation increased sharply as the MR severity increased. Overall, the underestimation by A2Ch HS-PISA (bias -52.0 ± 122.0 ml/s, P <.001; Figure 5E) is less than that by A4Ch HS-PISA (bias -65.5 ± 107.3 ml/s, P <.001; Figure 5F), while the underestimation by PLAX HS-PISA (bias - 77.6 ± 124.5 ml/s, P <.001; Figure 5D) was the largest.

**Figure 5.**
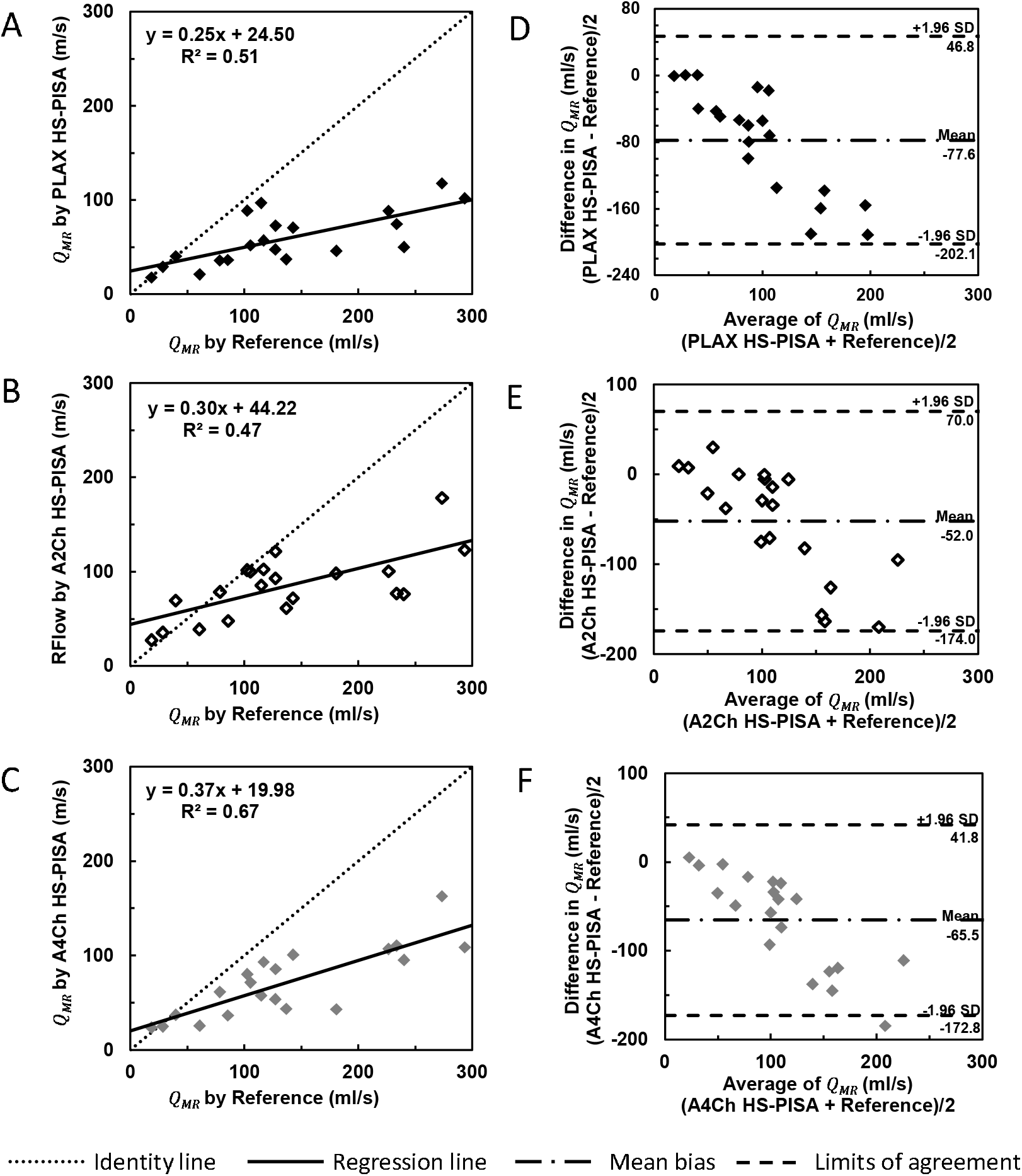
Linear regression plots showing correlations between *Q*_*MR*_ by reference, and *Q*_*MR*_ by PLAX HS-PISA (A), A2Ch HS-PISA (B), and A4Ch HS-PISA (C). Bland-Altman plots showing the *Q*_*MR*_ measurements bias by PLAX HS-PISA (D), A2Ch HS-PISA (E), and A4Ch HS-PISA (F).

### Measurement of regurgitant volume *RV*_*MV*_ : Peak PISA vs. Integrated PISA

The regurgitant volumes obtained with peak PISA and integrated PISA were compared with the reference values using Bland-Altman analysis (Table 2). The integrated PISA provided better agreement than the peak PISA for 3D-PISA (peak PISA bias: -12.7 ± 7.8 ml, integrated PISA bias: -2.1 ± 5.3 ml), HE-PISA (peak PISA bias: -20.5 ± 17.8 ml, integrated PISA bias: -12.8 ± 14.4 ml), HC-PISA (peak PISA bias: - 13.0 ± 18.7 ml, integrated PISA bias: -1.4 ± 14.5 ml), PLAX HS-PISA (peak PISA bias: -27.7 ± 25.2 ml, integrated PISA bias: -20.9 ± 23.8 ml), A2ch HS-PISA (peak PISA bias: -21.5 ± 30.2 ml, integrated PISA bias: -11.7 ± 25.3 ml), and A4ch HS-PISA (peak PISA bias: -22.2 ± 26.3 ml, integrated PISA bias: -16.3 ± 21.3 ml). Among different PISA methods, the accuracy in *RV*_*MV*_ quantification was higher using 3D-PISA, followed by multiplane 2D-PISA (HE-PISA and HC-PISA), and lastly single plane 2D-PISA (PLAX, A2ch, and A4ch HS-PISA).

**Table 2.** Results of Bland-Altman analysis for *RV*_*MV*_ quantification using both peak and integrated PISA

| PISA Method | Peak PISA |  | Integrated PISA |  |
| --- | --- | --- | --- | --- |
|  | Mean bias (ml) | Limits of agreement (ml) | Mean bias (ml) | Limits of agreement (ml) |
| 3D-PISA | -12.7 | [-20.4,-4.9] | -2.1 | [-7.5,3.2] |
| HE-PISA | -20.5 | [-38.3,-2.7] | -12.8 | [-27.2,1.6] |
| HC-PISA | -13.0 | [-31.7,5.7] | -1.4 | [-15.9,13.1] |
| PLAX HS-PISA | -27.7 | [-52.9,-2.5] | -20.9 | [-44.6,2.9] |
| A2ch HS-PISA | -21.5 | [-51.8,8.7] | -11.7 | [-36.9,13.6] |
| A4ch HS-PISA | -22.2 | [-48.5,4.1] | -16.3 | [-37.6,5.0] |

### Interobserver and Intraobserver Variability

A second interpreter performed blinded repeated measurements of a subset (20%) of the FMR models to assess interobserver variability. Good interobserver agreement for *RV*_*MV*_ quantification was shown, with an intraclass correlation coefficient of 0.95.

## DISCUSSION

### Quantification of PISA contour area

The accurate estimation of PISA contour area has always been the central issue of the PISA method. In 2D PISA, the area of PISA contour was not obtained directly, but was calculated based on certain assumptions of PISA contour shape. The conventional HS-PISA assumes a symmetric round orifice with hemispherical PISA contour. While this might be a reasonable assumption for degenerative MR, it does not hold for FMR, where an elongated and even curved orifice is typically observed along the leaflet coaptation. In FMR, the PISA of multiple jets merges and forms an extended and irregular contour [6, 34]. The conventional HS-PISA can only cover a relatively small portion of the 3D-PISA contour (Figure 6a), resulting in a significant MR underestimation. In all three HS-PISA (PLAX, A2ch and A4ch), we found that the MR underestimation was significant, especially for severe MR, which is consistent with previous studies [6, 20]. In order to improve the conventional single plane HS-PISA, various modified 2D PISA methods have been used [6, 12, 13, 34], which typically require measurements of the width (along the short axis), the length (along the long axis), and the vertical radius for PISA contour on multiple 2D planes. The HE-PISA assumes a hemiellipsoidal PISA contour, therefore is able to cover a bigger portion of the PISA (Figure 6b). However, for an asymmetric and curved orifice, HE-PISA would still miss the PISA contour towards the ends that extend along the anterolateral and posteromedial regions. The HC-PISA assumes a hemicylindrical PISA contour, which can include more area towards the ends along the long axis, especially with the inclusion of the semicircles on both ends (Figure 6c). Overall, our results showed that both HE-PISA and HC-PISA improved the quantification of *Q*_*MV*_ over the conventional HS-PISA, with a slight higher accuracy in HC-PISA.

**Figure 6.**
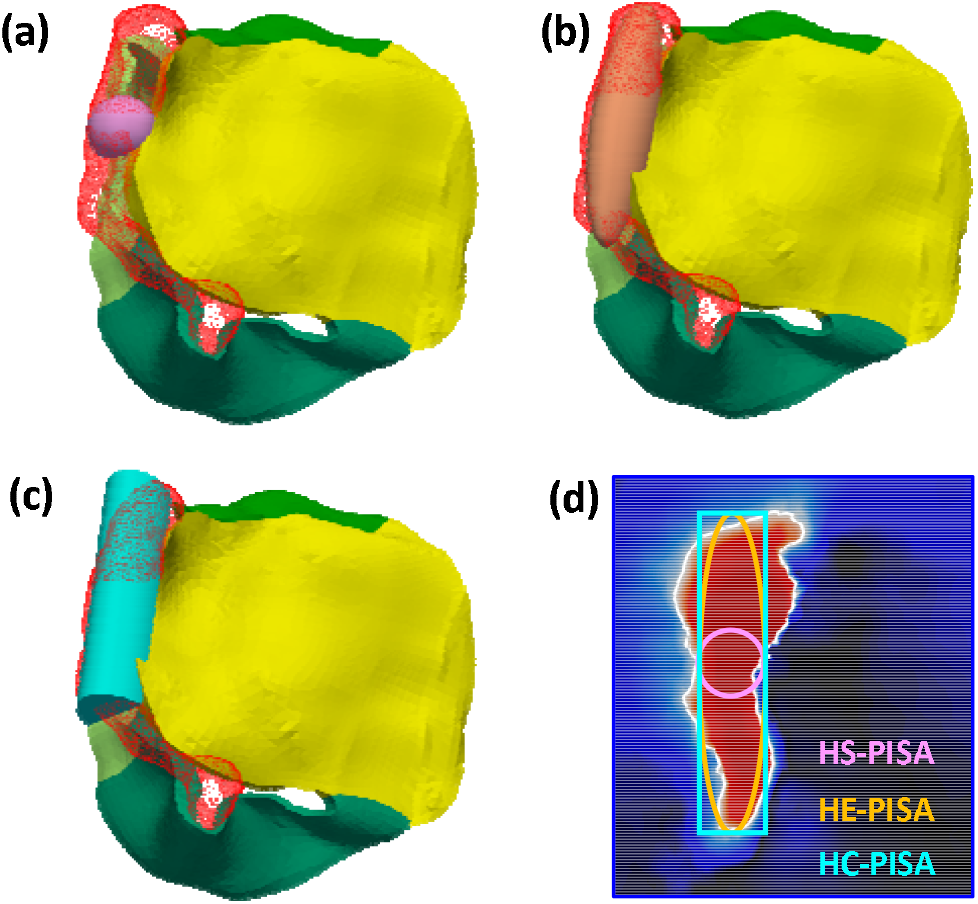
Comparison between 3D-PISA contour (red wireframe) and PISA contour calculated using HS-PISA (a, purple), HE-PISA (b, orange), and HC-PISA (c, cyan). In addition, (d) shows the comparison on short-axis view at PISA base, where the solid line denotes the base of the 3D-PISA. Purple circle, orange ellipse and cyan rectangular lines denote the base of HS-, HE- and HC-PISA, respectively.

Unlike 2D-PISA, 3D-PISA reconstructs the PISA contour from the 3D flow field, and measures the PISA area directly, without making any geometrical assumptions of the PISA contour. Therefore, despite the time consuming offline post processing [3], 3D-PISA overcomes the biggest limitation of the 2D PISA, and has attracted more attention recently [16, 35, 36]. However, it should be noted that the 3D-PISA still underestimated the true-PISA area (Figure 2d) due to the Doppler angle effect [2, 28-30]. While this underestimation may be small near the top of PISA; where the flow is nearly parallel to the ultrasound beam, it could be large near the PISA base; where the lateral flow forms a large angle with the ultrasound beam. Although the Doppler angle effect in PISA method has been well recognized, the magnitude of the associated underestimation has not been well defined in previous in vitro or clinical studies [2], due to the fact that true-PISA cannot be obtained. With both the true and the Doppler velocity field obtained from our computer models, we were able to quantify this error by comparing *S*_*PISA*_ of 3D-PISA to true-PISA, which showed a relative underestimation of 35.2 % ± 13.1 % in *S*_*PISA*_ obtained using 3D-PISA.

### Quantification of the instantaneous regurgitant flow rate *Q*_*MR*_

Based on fundamental fluid mechanics principles, the regurgitant flow rate through PISA is *Q*_*MR*_ = *∫*_*s*_ *V*_*n*_ *dS*, where *V*_*n*_ is the normal component of the true velocity across PISA. The simplified expression *Q*_*MR*_ = *V*_*a*_ *S*_*PISA*_ used in the PISA methods will therefore only be accurate when *V*_*n*_ = *V*_*a*_, which is not always the case (Figure 7). Due to the Doppler angle effect, the aliasing velocity detected by echo *V*_*a*_ =*V* cos *θ* underestimates the true velocity *V*, where *θ* is the “Doppler angle” between the direction of flow and the ultrasound beam. However, as commented in Ref. [2], the relatively large underestimation caused by the Doppler angle effect seems to be balanced by a systematic overestimation from some other parameter, resulting relatively small underestimation in 3D-PISA. In this study, we found that this systematic overestimation is likely due to the second angle (overlooked in most studies of PISA method): the “flow direction angle”, defined as the angle between the flow direction across PISA and the normal direction of PISA. Since only the normal component *V*_*n*_ = *V* cos *γ* will contribute to the flow rate across PISA, *γ* would lead to overestimation in *Q*_*MR*_. Indeed, for true-PISA which eliminated the errors due to Doppler angle effect, a consistent overestimation in *Q*_*MR*_ (bias 32.3 ± 35.2 ml/s, P <.001; Figure 3C) was observed due to the error caused by *γ*.

**Figure 7.**
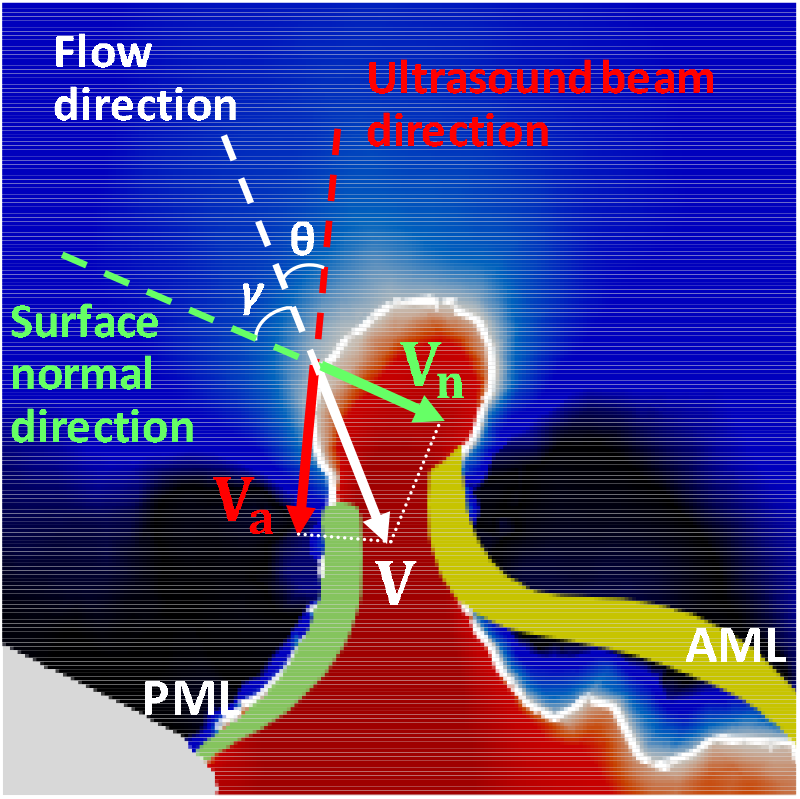
Schematic showing the flow velocity *V* across PISA, the component *V*_*n*_ normal to PISA, and the component *V*_*a*_ detected by echo. The angle *θ* is the Doppler angle between flow and the ultrasound beam direction, and *γ* is the flow direction angle between the flow and the surface normal direction of PISA.

As 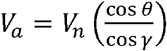, the underestimation due to the Doppler angle *θ*, and the overestimation due to the flow direction angle γ compensate each other. When *θ* and γ are close, *Q*_*MR*_ obtained using PISA method could be close to the reference value, as long as *S*_*PISA*_ is accurately captured. This could be one of the main reasons that multiple studies have reported relatively accurate quantification of MR using 3D-PISA despite the Doppler angle effect [15, 16].

### Quantification of regurgitant volume *RV*_*MV*_ : Peak PISA vs. Integrated PISA

Finally, for *RV*_*MV*_ quantification, an additional source of error is due to the dynamic feature of the regurgitant orifice, especially in FMR [8, 37]. The accurate regurgitant flow volume *RV*_*MV*_ is simply a time integral of *Q*_*MR*_, which can be approximated as a sum over all of the *n* frames during systole: 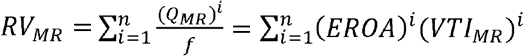, where *f* is the color Doppler frame rate. This suggests that the peak PISA obtained using the measurement at a single frame is only accurate when EROA is constant over systole, and the “integrated PISA” using multiple frame measurement over systole is more accurate. It should be noted that different ways of processing multiple frames have been reported [7, 18, 19], and one should be careful to perform “integrated PISA” properly. If 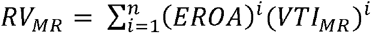 is used, (*EROA*)^*i*^ at frame *i* should be calculated using (*Q*_*MR*_)^*i*^ /(*V*_*MR*_)^*i*^ from that same frame, and partial (*VTI*)^*i*^ between two frames should be used instead of the complete *VTI*_*MR*_ over the entire systole. In particular, the product of 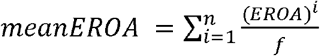 [19], and the complete *VTI*_*MR*_ will not yield accurate results. In fact, considering that (*V*_*MR*_)^*i*^ at each frame and partial (*VTI*)^*i*^ are not easily accessible in clinical practice, 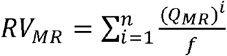 [7, 18] is recommended since it only requires a simple sum of (*Q*_*MR*_)^*i*^ over systole, without even the requirement of CWD measurements for *VTI*_*MR*_ nor the calculation of EROA.

### Limitations

There are some limitations in this study. The computer FMR models employed the same baseline loading conditions for the cardiac wall movement, and focused on a type IIIb lesion with restricted leaflet motion. Other types of FMR such as type I MR with normal leaflet motion have not been investigated, which warrants a future study. Additionally, virtual echo represents an ideal measurement with optimal positioning and angle of the echo probe, hence certain sources of error for clinical echo such as reverberation artifacts and shadowing were not incorporated.

## CONCLUSIONS

We evaluated the accuracy of different PISA methods by comparing virtual echo results with reference values obtained from computer FMR models. These computer models enabled us to identify and quantify different sources of errors in MR quantification. It was found that for 2D PISA, the error was mostly due to the quantification of the PISA area. Conventional HS-PISA significantly underestimated FMR with elongated orifice, while HE- and HC-PISA could lead to improvements using measurements of PISA dimensions on multiple 2D planes. For 3D-PISA, although the actual PISA contour is captured rather than calculated, the underestimation due to “Doppler angle” *θ* and overestimation due to “flow direction angle” γ still leads to uncertainty in *Q*_*MR*_ quantification. Finally, while integrated PISA is more accurate than peak PISA for *RV*_*MR*_ quantification, it needs to be done properly in order to correctly capture the dynamic feature of MR. In specific, 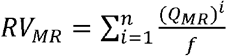 using a simple sum of *Q*_*MR*_ is the most accurate and convenient way for *RV*_*MR*_ quantification.

## Data Availability

All the data was included in the paper

## ABBREVIATIONS AND ACRONYMS

MR: mitral regurgitation
Echo: echocardiography
FMR: functional mitral regurgitation
PISA: proximal isovelocity surface area
EROA: effective regurgitant orifice area
RV: regurgitant volume
RF: regurgitant fraction
FSI: fluid-structure interaction
PM: papillary muscle
AV: aortic valve
MV: mitral valve
AML: anterior mitral leaflet
PML: posterior mitral leaflet
LV: left ventricle
LA: left atrium
SV: stroke volume
HE: hemiellipsoidal
HC: hemicylindrical
HS: hemispherical
PLAX: parasternal long-axis
A2ch: apical 2-chamber
A4ch: apical 4-chamber
CWD: continuous wave Doppler
VTI: velocity time integral

## ACKNOWLEDGEMENTS

Dr. Tongran Qin was in part supported by the American Heart Association (AHA) Postdoctoral Fellowship 19POST34450161. Dr. Andrés Caballero was in part supported by a Fulbright-Colciencias Fellowship.

## Notes

### Competing Interest Statement

Dr. Wei Sun is a co-founder and serves as the Chief Scientific Advisor of Dura Biotech. He has received compensation and owns equity in the company. Dr. Rebecca Hahn is a speaker for Abbott Vascular, Boston Scientific, Edwards Lifesciences, Philips Healthcare, and on the advisory board/consultant for Edwards Lifesciences, Gore & Associates, Medtronic, and Navigate. The remaining authors have nothing to disclose.

### Author Declarations

IRB-approved protocols were obtained from Georgia Tech Institutional Review Board and Columbia University Institutional Review Board.

